# Masks in a Post COVID-19 World: A Better Alternative to Curtailing Influenza?

**DOI:** 10.1101/2021.07.03.21259943

**Authors:** Henri Froese, Angel G. A. Prempeh

**Affiliations:** Goethe-University Frankfurt am Main, Saginaw Valley State University

## Abstract

Over the course of the coronavirus pandemic, it has become apparent that non-pharmaceutical interventions such as masks and social distancing are of great help in mitigating the transmission of airborne infectious diseases. Additionally, data from respiratory specimen analysis from the past year show that current mask mandates established for COVID-19 have inadvertently reduced the rates of other respiratory diseases, including influenza. Thus, the question arises as to whether comparatively mild measures should be kept in place after the pandemic to reduce the impact of influenza. In this study, we employed a series of differential equations to simulate past influenza seasons, assuming people wore face masks. This was achieved by introducing a variable to account for the efficacy and prevalence of masks and then analyzing its impact on influenza transmission rate in an SEIR model fit to the actual past seasons. We then compared influenza rates in this hypothetical scenario with the actual rates over the seasons. Our results show that several combinations of mask efficacy and prevalence can significantly reduce the burden of seasonal influenza. Particularly, our simulations suggest that a minority of individuals wearing masks greatly reduce the number of influenza infections. Considering the efficacy rates of masks and the relatively insignificant monetary cost, we highlight that it may be a viable alternative or complement to influenza vaccinations. We conclude with a brief discussion of our results and other practical aspects

## 1 Introduction

In March 2020, the WHO officially declared COVID-19 a global pandemic as it extended beyond borders and reached out to various parts of the world([1]). The spread of the virus has halted several activities and has placed uncertainty on future events. Scientists and researchers have recommended safety measures like social distancing, wearing of masks and quarantines to reduce infection rates, or “flatten the curve” ([2]). In the advent of this new reality, current analysis of respiratory specimens from 2018-20 in Hong Kong indicate that rates of other respiratory pathogens such as Respiratory Syncytial Virus and influenza are decreasing with increased mask-wearing([3]). This is not unique to Hong Kong,([4]) presents data from the United States, Australia, Chile, and South Africa also showing significantly reduced rates of influenza following the widespread adoption of non-pharmaceutical interventions such as masks. To gain an in-depth and quantitative understanding of the impact of face masks on the reduction in influenza activity, we simulate how past influenza seasons 2010/11 to 2018/19 would have played out *had people worn masks*.

The simulations were developed using deterministic compartmental models with the incorporation of variables to account for the impact of masks. Using publicly available influenza infection data for the past seasons from the CDC, the model for each season (2010/11 to 2018/19) was calibrated. We then simulate the seasons factoring in different scenarios of mask prevalence as well as inward-outward efficacy. The results of these counterfactual simulations are described and compared in section 3 and discussed in section 4.

## 2 Methods

In this section, parameters involved in the deterministic Susceptible-Exposed-Infected-Recovered model(SEIR) was described. Then, the mathematical underlinings of the mask parameter were evaluated.

### 2.1 SEIR model and parameters

The SEIR-model with time-dependent transmission rate is described by the following equations:

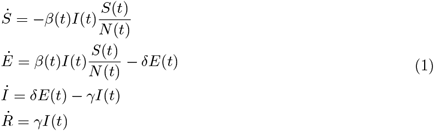

Where *S*(*t*), *E*(*t*), *I*(*t*), *R*(*t*) are the number of susceptible, exposed (infected, not infectious yet), infected, and recovered individuals at a given time *t*. Since the flu fatality rates are insignificant in relation to the total population([5]),deaths from the flu, and unrelated births and deaths were disregarded. The parameter *δ* describes the rate of progression from exposed to infected, that is, the inverse of the incubation period. The parameter *γ* is the rate of progression from infected to recovered or the inverse of the generation time.

Transmission rate *β*(*t*) is described as the number of contacts an infected individual has per timestep, multiplied by the probability of disease transmission in a contact. Thus, as only the fraction 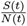 of the population can be infected, every infected infects 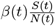 individuals per timestep. In regards to influenza, all parameters of the SEIR-model except the time-dependent transmission rate *β*(*t*) are publicly available via CDC data ^1^. Hence, we estimate *β*(*t*) by fitting the model to the scaled past infection data.

To account for mask usage, a simplified version of the model used in [6] was adopted.

- *m*_pre_ ∈ [0, 1] is the mask prevalence, taken as the proportion of contacts in which an individual wears a mask. We assume that infection status does not affect mask-wearing behavior.
- 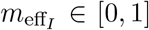 is the efficacy of mask usage by the infected individual, i.e. the reduction of the chance of infection when only the infected wears a mask.
- 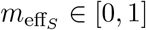 is the efficacy of mask usage by the susceptible individual, i.e. the reduction of the chance of infection when only the susceptible wears a mask.

Consequently, we assume that the reduction of the chance of infection when both individuals in a contact wear a mask is 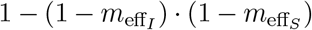. For example, if the outward efficacy is 0.7 and the inward efficacy is 0.9, then the infection only happens in 3% of contacts where both individuals wear a mask. We combine the parameters to define the *mask impact m* ∈ [0, 1], the *proportion of contacts in which masks prevent an infection* given the three parameters listed above. That is the sum of the proportions of contacts prevented if both individuals wear masks, only the infectious individual wears a mask, only the susceptible individual wears a mask, or no one wears a mask, leading to the following formula that sums these four cases up:

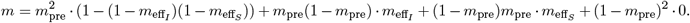

To incorporate *m* (the proportion of contacts prevented through mask usage) into the model, note that without masks, every infected infects 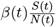 individuals per timestep – thus, with masks, this changes to 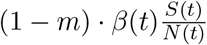. We get the following model:

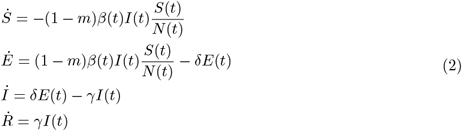

We will now look at the data used to fit *β*(*t*) for this model to past flu seasons (without masks, i.e. with *m* = 0).

### 2.2 Infection data

The CDC FluView application^2^ provides weekly number of positive flu tests (we do not separate between different strains) in public health and clinical laboratories for seasons 2010/11 to 2018/19. To extrapolate from the weekly number of positive flu tests to the weekly number of infected individuals, we additionally use the CDC’s estimated total number of infections per season^3^.

For any season, let *P*_*i*_ be the number of positive flu tests in week *i*. Let *T* be the total number of infections for the season. We assume that the number of positive tests is proportional to the actual number of infected *I*_*i*_ that we are interested in, that is, *I*_*i*_ = *λP*_*i*_, for all weeks *i*, for some fixed (per season) scaling factor *λ >* 0. As infections persist on average for one week (see the description of known parameters in 2.3), we get that the sum of infected per week over all weeks is (approximately) the total number of infections for the season: ∑_*i*_ *I*_*i*_ = *T*. We thus have ∑_*i*_ *λP*_*i*_ = ∑_*i*_ *I*_*i*_ = *T*, and can solve for the season’s scaling factor with

For each season, we calculate the scaling factor *λ* and use it to scale the CDC data.

### 2.3 Beta estimation from infection data

To estimate the time-dependent transmission rate, we fit a seasonal function of the form

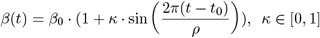

to the scaled data for each season, similar to approaches in e.g. [7], [8].

Timesteps *t* are in weeks. The incubation period and generation time are adapted from [9], yielding *γ* = 1.0 (infections last one week) and 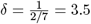 (incubation period of two days).

Least-Squares fitting using the LMFIT Python library ([10]) yields good fits on all seasons. See figure 1 for a visualization of the data and the achieved fit.

**Figure 1:**
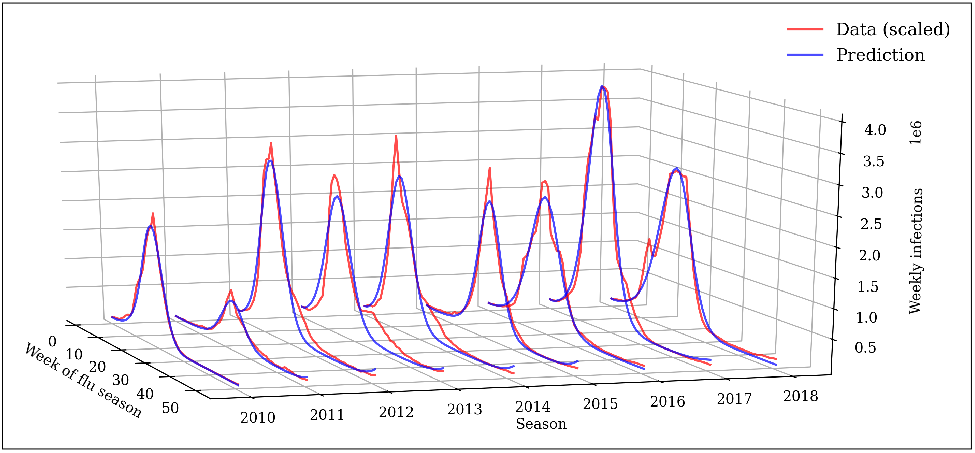
Results of transmission rate fitting to data of past flu seasons: Actual infection data (scaled as outlined in 2.2) and prediction for influenza seasons 2010/11 to 2018/19.

**Figure 2:**
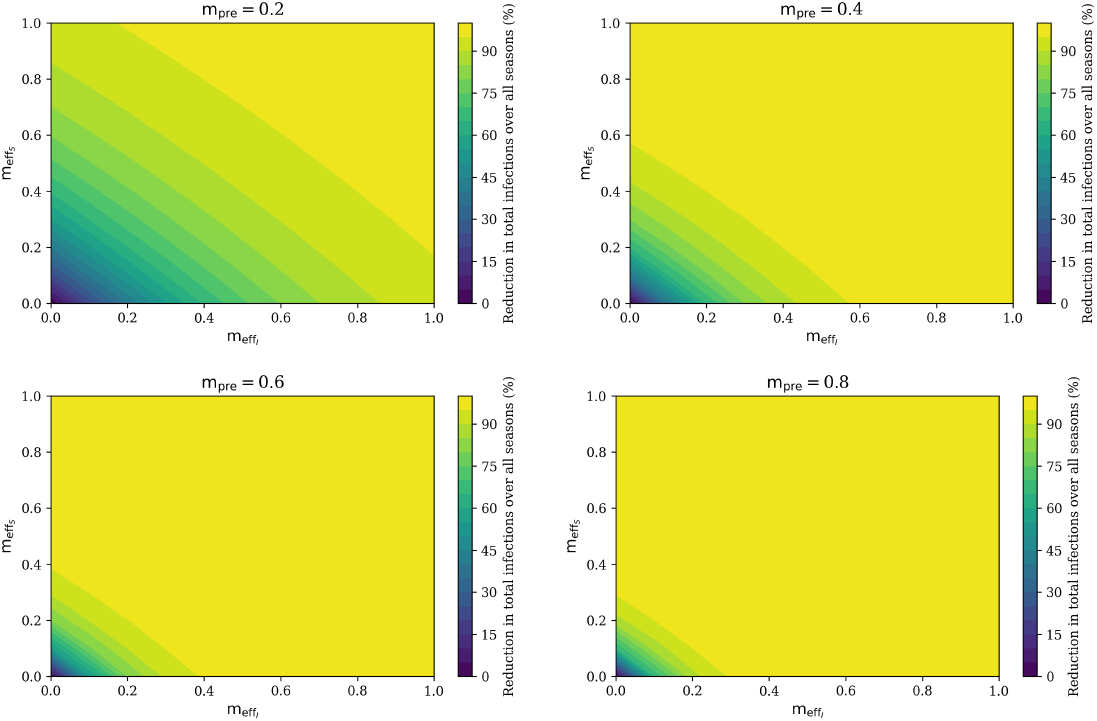
(Reduction of) total infections with mask usage for different choices of mask prevalence *m*_pre_, reduction in chance of infection when the infectious individual wears a mask 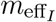, and reduction in chance of infection when the susceptible individual wears a mask 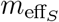.

## 3 Results

We simulate the past influenza seasons with the estimated transmission rate *β*(*t*) and compare the outcome with and without masks. As evidenced in [11] and [12], mask efficacy is highly uncertain. Therefore, different combinations of mask prevalence and outward and inward efficacy are implemented.

Findings show that when mask prevalence is high, say over 0.6, low efficacies (for example caused by masks worn too long, loose-fitting, etc.) are sufficient to fully contain the flu. And even with low prevalence, inward and outward efficacies of around 0.5 prevent over 70% of infections. As such efficacies might be likely as described in [6], it seems as though a minority of disciplined mask-wearers is sufficient to prevent most infections.

Mask use during the Coronavirus pandemic in the US ranged from 50% to 70% from May to December 2020^4^. We believe it is unlikely that similar acceptance rates can be achieved after the Coronavirus pandemic without a mask mandate. As evidenced by the significant reduction in infections for even low efficacies when prevalence is high, a mask mandate would certainly be highly effective in containing the flu. Figure 3 shows detailed results for all seasons for two scenarios we deem the most relevant: the ‘mask mandate’ scenario with a prevalence of 0.5 and outward and inward efficacies of 0.35, and the ‘masks suggested’ scenario with a prevalence of 0.2, with outward and inward efficacies of 0.45. We thus err on the lower range of efficacy estimates. Behind the assumed higher efficacy in the second scenario lies the assumption that individuals wearing masks without a mandate are more likely to wear high-quality, well-fitting masks.

**Figure 3:**
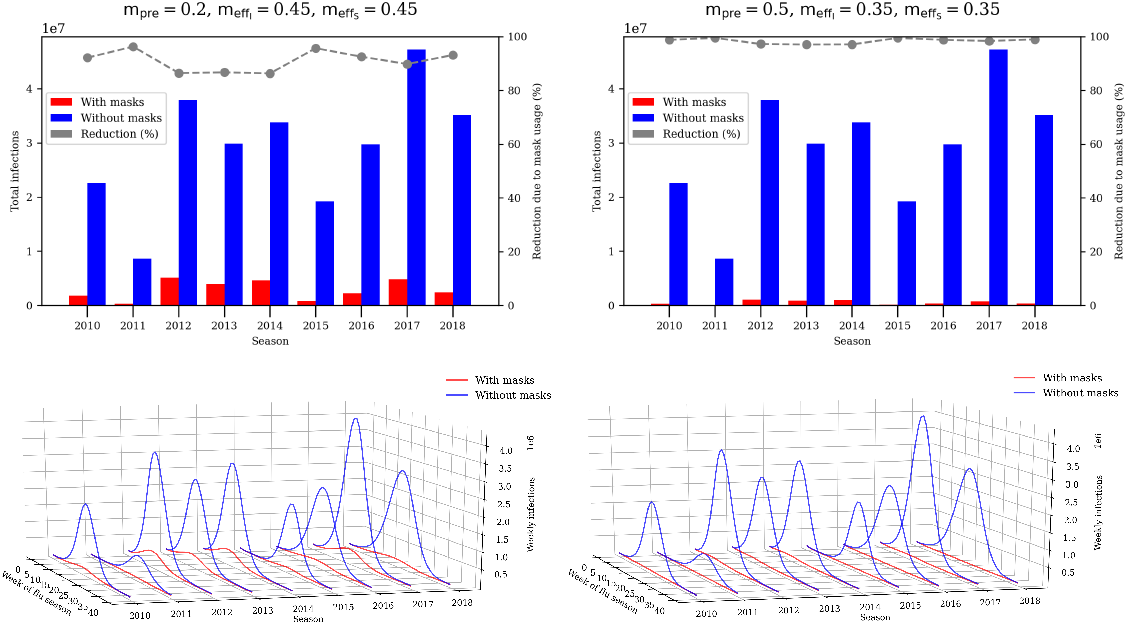
Weekly and total infections with mask usage by season, simulated for the ‘masks suggested’ scenario (left) and the ‘mask mandate’ scenario (right). The simulations how that in both scenarios, most infections are prevented in each season. The ‘mask mandate’ scenario succeeds in fully containing the flu in most seasons.

## 4 Discussion

Our simulations show that a relatively low mask prevalence of around 0.2 and assumed moderate inward and outward efficacy of 0.45 would have greatly reduced influenza infections by *>* 90% over several past seasons. Additionally, 0.5 mask prevalence combined with lower efficacies leads to *>* 95% reduction in influenza illnesses. Limitations of our approach include no stratification by age or contact scenario, significant uncertainties in mask use and efficacy, and disregard of other non-pharmaceutical interventions.

Currently, vaccinations are the prominent way to protect against influenza, having been available on a large scale since 1945 ([13]). Current influenza vaccination rates in the US are not high enough to provide herd immunity ([14]), and have averted around 15-20% of influenza illnesses over the seasons from 2011-12 to 2018-19 ^5^. While, vaccines have to be newly manufactured each season with significant R&D investments, the vaccines of course only have to be administered once while face masks would need to be worn continuously, which might be seen as more burdensome by the general population.

[15] estimates the economic burden of seasonal influenza in the United States at $6.3 -$25.3 billion. Assuming the economic cost scales linearly with the number of infections, a scenario in which at least 95% of infections are reduced (which includes both the ‘mask mandate’ and ‘masks suggested’ scenarios) saves $6.0 – $24.0 billion per season, at negligible cost. Similar to public opinion regarding potential health hazards such as smoking and driving without seatbelts shifting over time, and legislation being introduced, we can imagine the Coronavirus pandemic changing public (and expert) opinion towards everyday mask usage; then again, large parts of the population might be tired of wearing masks after the Coronavirus pandemic. If public opinion shifts and (at least) a minority of individuals wears masks, our simulations show that this would greatly reduce the significant burden of seasonal influenza, at a little monetary cost.

## Data Availability

The authors confirm that the data supporting the findings of this study are available within the article and its supplementary materials.

## Footnotes

1 https://www.cdc.gov/flu/about/burden/past-seasons.html

2 https://www.cdc.gov/flu/weekly/fluviewinteractive.htm

3 https://www.cdc.gov/flu/about/burden/past-seasons.html

4 https://covid19.healthdata.org/united-states-of-america?view=mask-usetab=trend

5 https://www.cdc.gov/flu/vaccines-work/past-burden-averted-est.html

## Notes

### Competing Interest Statement

The authors have declared no competing interest.

### Funding Statement

This research received no funding or grant from any funding agency in the public, commercial, or not-for-profit sectors.

### Author Declarations

Research does not involve human subjects and are not subjected to IRB approval or exemption

